# Specialty pharmacy disease screening and routine assessments for patients with rheumatoid arthritis and psoriasis

**DOI:** 10.1101/2022.06.06.22276034

**Authors:** Alana J. Dube, Kenneth L. McCall, Kirsten E. Stickney, Alycia Gelinas

## Abstract

**Background:** Rheumatoid arthritis and psoriasis are inflammatory diseases which require frequent monitoring to optimize therapy. Specialty pharmacists are in the unique position to assist in the screening and monitoring of patients with complex, chronic diseases.

**Objectives:** The study objective is to describe the impact of pharmacist screening services in two patient populations. In patients with rheumatoid arthritis, the goal is to describe outcome monitoring through disease severity, therapeutic switches, and adherence rates. In patients with psoriasis, the aim is to describe the utilization of a screening for psoriatic arthritis and the resulting number of potential referrals to medical providers.

**Methods:** The retrospective study patient population consisted of rheumatoid arthritis (RA) patients who filled one or more prescriptions at a specialty pharmacy between 8/22/2017 and 9/26/2018, and psoriasis patients who filled between 6/1/2021 to 9/1/2021. A Routine Assessment of Patient Index Data 3 (RAPID3) score was collected during a refill coordination call every three months throughout the 13-month period for RA patients, and a Psoriasis Epidemiology Screening Tool (PEST) scores reported throughout the stated timeline.^1,2^ Data was imported from the pharmacy’s electronic medical record into an Excel spreadsheet with each row representing a unique patient. Following data validation, descriptive statistics including means, standard deviations, and percentages were calculated to characterize the sample. Statistical significance was determined at an alpha of 0.05.

**Results:** Of the patients who had 4 assessments for RAPID3, the disease severity category significantly improved from assessment 1 to assessment 4 (p=0.021) when analyzed using a chi-square test. The RAPID3 assessment of patients with RA by pharmacists in a specialty setting identified responders (n=21, 25.6%) and stable patients (n=51, 63%), which reinforces current therapy, and non-responders (n=10, 12.2%), who may benefit from referral to their provider for reevaluation of their therapeutic plan. The PEST screening of patients with psoriasis by pharmacists in a specialty setting identified 11 of 32 patients (34%) who scored a 3 or higher and who may benefit from a referral to a rheumatologist for further assessment of psoriatic arthritis activity.

**Conclusion:** Specialty pharmacists are an essential part of ongoing assessment and management of patients with chronic inflammatory conditions such as rheumatoid arthritis and psoriasis. Screening and monitoring of patients by pharmacists can identify the need for referral to a medical provider.

**Summary Bullets:** What is already known about this subject?

Current guidelines for the treatment of rheumatoid arthritis recommend frequent monitoring and reassessment every three months until low disease activity or remission is achieved. Similar screening efforts in patients with psoriasis can help identify the nearly 30% of patients who have undiagnosed psoriatic arthritis. Clinical pharmacists in the specialty setting can assist with these screening initiatives to reduce disease severity and appropriately refer patients for further examination.

What this study adds.

This study demonstrates the ability of clinical specialty pharmacists to administer validated screening tools used in chronic inflammatory disease states to improve patient outcomes.

**Disclosures:** The authors of this study have no possible financial or personal relationships with commercial entities to disclose that may have a direct or indirect interest in the matter of this study.

**Funding source:** None.

## INTRODUCTION

Specialty pharmacists are in the unique position to assist in screening initiatives and disease state management in patients with complex auto-immune disorders such as rheumatoid arthritis (RA) and psoriasis (PsO). Extensive knowledge of biologic therapies and correlating conditions provide these health professionals with the tools needed to administer assessments such as the Routine Assessment of Patient Index Data 3 (RAPID3) for RA and The Psoriatic Epidemiology Screening Tool (PEST) for psoriasis.^3,4^ Additionally, pharmacists receive the training and qualifications needed to appropriately tailor drug regimens in an effort to optimize treatment of these progressive inflammatory conditions.

Rheumatoid arthritis is an autoimmune disorder of the joints characterized by chronic inflammation, pain, and a significant impact on quality of life.^5^ Data from the Rheumatology Informatics System for Effectiveness (RISE) registry showed that nearly 50% of treated patients still had moderate to high disease activity and did not change their therapy over the next year, highlighting the need for close monitoring and intervention as a key strategy of disease management to prevent further disease progression^.6^ The 2021 American College of Rheumatology (ACR) guidelines recommend assessing and adjusting therapy at least every 3 months until disease remission or low activity is achieved.^7^ This method has been associated with improved patient outcomes, with approximately 60% of patients achieving remission when compared to conventional care.^8,9^ These recommendations largely focus on the importance of shared-decision making between the patient and provider and maximizing the treat-to-target approach through use of disease activity assessments such as DAS28, CDAI, and RAPID3.^10,11^

RAPID3 is a disease activity index used to measure disease severity in RA patients through assessment of the 3 core data set measures: function, pain, and patient global estimate of status. Scores can range on a scale of 0 to 30, with higher scores indicating worse disease severity. Strengths of RAPID3 include its simplicity, ability to be conducted in a timely manner, and endorsement from the ACR as a reliable tool for standard monitoring initiatives in this patient population.^12^ The direct correlation to more detailed disease assessments such as the Disease Activity Score 28 (DAS28) and Clinical Disease Activity Index (CDAI) has been demonstrated through numerous clinical trials and high prevalence of use in clinical care settings.^4^

Psoriasis is another chronic and relapsing inflammatory condition characterized by plaques and scales on the skin as a result of keratinocyte hyperproliferation.^13^ Up to 42% percent of patients with psoriasis will go on to develop psoriatic arthritis, with an alarming 30% of these patients left undiagnosed with a progressive joint disease.^9^ Additionally, as many as 47% of psoriatic arthritis patients develop erosive joint damage within 2 years of symptom onset, validating the recommendation to screen patients at every visit from the joint American Academy of Dermatology-National Psoriasis Foundation (AAD-NPF) guidelines to encourage early detection and referral to rheumatologist.^14^

PEST is a 5-item questionnaire, which includes a joint diagram to help identify comorbid psoriatic arthritis in patients with psoriasis. The scale ranges from 0 to 5, with higher scores associated with increased psoriatic arthritis activity, and scores ≥ 3 suggesting a benefit in referral to a rheumatologist. The goal of the screening is to manage pain and prevent further joint damage that may otherwise go untreated.^15^ This assessment is validated for use by general practitioners and dermatologists and has been identified as one of the easiest psoriatic arthritis screening tools for patients to complete and for physicians to score.^16^ In addition to worsening disease in the affected areas such as the joints and skin, untreated or sub-optimally treated inflammation has the potential to threaten the health of other bodily systems including cardiovascular, renal, and ocular. When these inflammatory conditions are not treated adequately, there can be increased rates of diabetes, heart disease, chronic kidney disease, COPD, and elevated risk of many different types of cancers.^17^

Provider shortages and increasing workloads within specialty practices such as rheumatology and dermatology, are current barriers to the proper screening and treatment recommended by the American College of Rheumatology, and the American Academy of Dermatology, respectively.^18,19^ These patient populations require frequent monitoring, reassessment, and therapy adjustments. Pharmacists have the ability to play an important role in maximizing interprofessional collaboration within the healthcare team to fully adapt guideline recommendations and improve patient outcomes. The evidence of pharmacist involvement in screening initiatives has been shown through numerous other disease states, including peripheral artery disease, COPD, and osteoarthritis.^20,21,22^ There are currently no studies published which demonstrate the benefits of specialty pharmacist-administered screening initiatives to assess the severity and disease control of RA using RAPID3.

## METHODS

The setting of this study is a specialty pharmacy licensed in multiple states and with dual accreditation by the URAC and the Accreditation Commission for Health Care. Pharmacists in the practice setting have earned the Certified Specialty Pharmacist credential from the National Association of Specialty Pharmacy. The retrospective study patient population consisted of rheumatoid arthritis (RA) patients who filled one or more prescriptions at a specialty pharmacy between 8/22/2017 and 9/26/2018, and psoriasis patients who filled between 6/1/2021 to 9/1/2021. A Routine Assessment of Patient Index Data 3 (RAPID3) score was collected during a refill coordination call every three months throughout the 13-month period for RA patients, and a Psoriasis Epidemiology Screening Tool (PEST) scores reported throughout the stated timeline.^1,2^ Manual retrospective chart reviews and reports generated from the pharmacy’s electronic medical record system were previously used to collect information.

Additional data reviewed at the time of survey assessment included biologic medication, fill dates, prescriber demographics, ICD-10 code for medical diagnosis, and medication history, including drug, dose, and frequency. The primary endpoints consist of describing pharmacist involvement in screening services and routine assessments in three major areas: disease activity, therapeutic changes, and potential referrals to medical providers. Adherence rates, average pain scores, and overall function and well-being may be reviewed as information that pharmacists are able to collect and provide to physicians but will not be directly analyzed within this descriptive review. This study will be describing data that was previously collected, de-identified, and analyzed using Microsoft Excel and R version 4.1.1.

## STATISTICAL ANALYSIS

Data was imported from the pharmacy’s electronic medical record into an Excel spreadsheet with each row representing a unique patient. Data was encoded numerically when needed. For quality assurance, the data was cleaned to ensure that there were no transcription errors. Following data validation, descriptive statistics including means, standard deviations, and percentages were calculated to characterize the sample.

## RESULTS

As shown in Table 1, a total of 127 patients were given the RAPID3 ≥ 1 time during the study period. The overall mean age of patients was 52 years, with a gender distribution of 76% female and 24% male.

**Table 1:** Baseline demographics among RAPID3 participants

| Parameter | Male | Female | Overall |
| --- | --- | --- | --- |
| Number of participants | 30 (23%) | 98 (77%) | 128 |
| Total number of survey responses | 101 (26%) | 288 (74%) | 389 |
| Average age in years | 54.5 | 51.2 | 52.2 |
| Average years with RA | 8.1 | 12.3 | 11.2 |

As shown in Figure 1, the mean RAPID3 score was 6.8 at the third measurement with overall average composite RAPID3 scores trending from 10.0 (moderate severity) at assessment #1, to 5.2 (low severity) at assessment #6.

**Figure 1:**
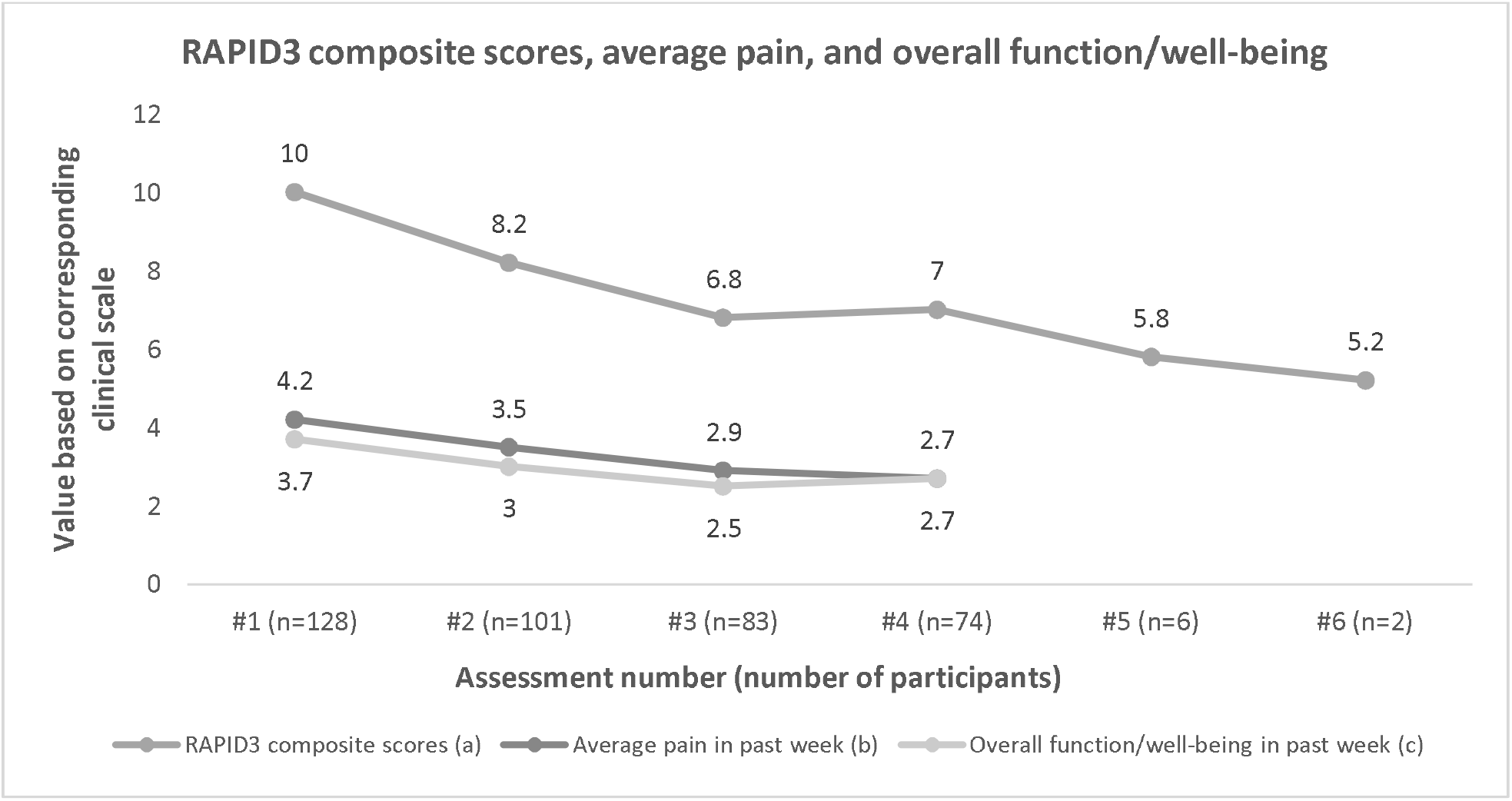
RAPID3 composite scores, average pain, and overall function/well-being ^a^RAPID 3 is a composite score of physical function, pain, and patient global estimate on a 0 to 30 scale. ^b^Pain was assessed on a 10-point scale. ^c^Overall function/well-being was assessed on a scale from 0 to 5, with lower numbers reflecting improved function.

As shown in Figure 2, a total of 11 patients had therapeutic switches during the 13-month assessment period. One patient had 2 switches and another patient had a total of 3 switches throughout the given timeline.

**Figure 2:**
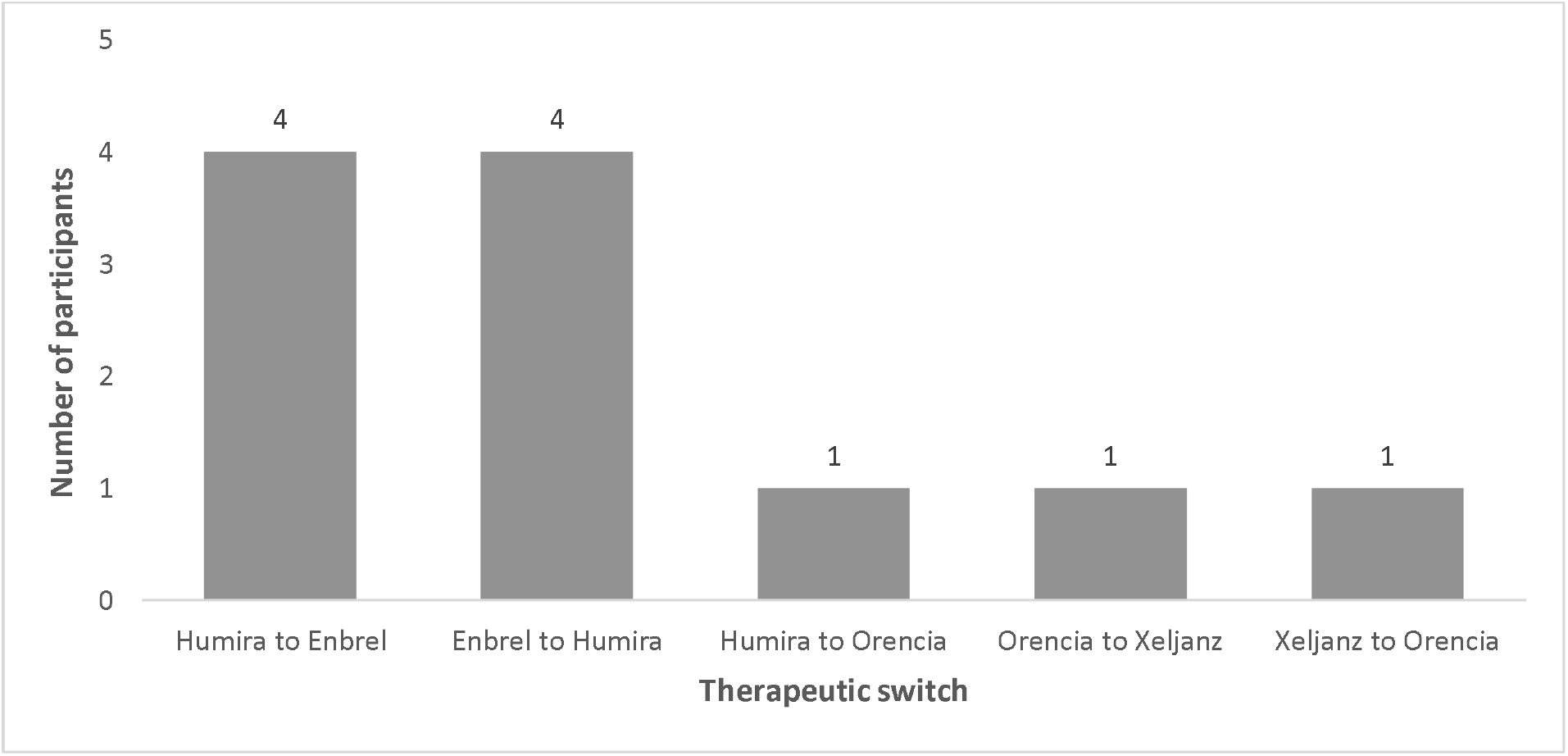
Therapeutic switches among rheumatoid arthritis patients over 13-month assessment period

Of the patients who had 4 assessments for RAPID3, the disease severity category significantly improved from assessment 1 to assessment 4 (p=0.021) when analyzed using a chi-square test. Although there was a statistically significant overall improvement in disease severity category, not all patients improved; 21 patients were classified as responders, improving by at least one disease severity category. 51 patients had no change in disease severity category (Of note, 25% of patients were classified to be in remission at assessment #1), and 10 patients were non-responders, worsening by a disease severity category. The proportion of responders was determined to be 25.6% (n=21) and proportion of non-responders was 12.2% (n=10). The RAPID3 assessment of patients with RA by pharmacists in a specialty setting identified responders (n=21, 25.6%) and stable patients (n=51, 63%), which reinforces current therapy, and non-responders (n=10, 12.2%), who may benefit from referral to their provider for reevaluation of their therapeutic plan. In addition, a descriptive examination of average pain levels on a scale from 0 to 10 showed a trend down from 4.2 at assessment 1, to 2.7 at assessment 4. Similarly, overall function and well-being averaged 3.7 upon initial assessment, and 2.73 at the final assessment with lower numbers reflecting improved function. Of note, a large proportion of the 11 patients with therapeutic switches (n=7, 70%) were among those who scored a 12 or higher on the RAPID3 assessment, an indicator of severe disease activity. This data would suggest that patients were seen for follow-up by their rheumatologist and appropriately changed to an alternative therapy in an attempt to treat-to-target as current ACR guidelines recommend.

The mean age of PEST screen participants was 48 years old, (45.7 for females and 50.6 for males) with a standard deviation of 12.95 years among both genders. The study was comprised of 53% females (n=17) and 47% males (n=15). Frequency of drug regimens used was as follows: Cosentyx (secukinumab) 3, Humira (adalimumab) 7, Otezla (premilast) 6, Otezla + Enbrel (apremilast + etanercept) 1, Skyrizi (risankizumab) 4, Stelara (ustekinumab) 4, Taltz (ixekizumab) 6, Tremfya (guselkumab) 1. Figure 3 shows the PEST scores by patient count. A total of 11 patients (34%) screened positive (score ≥ 3) with the PEST screening tool, indicating psoriatic arthritis activity and the opportunity for referral to a rheumatologist.

**Figure 3:**
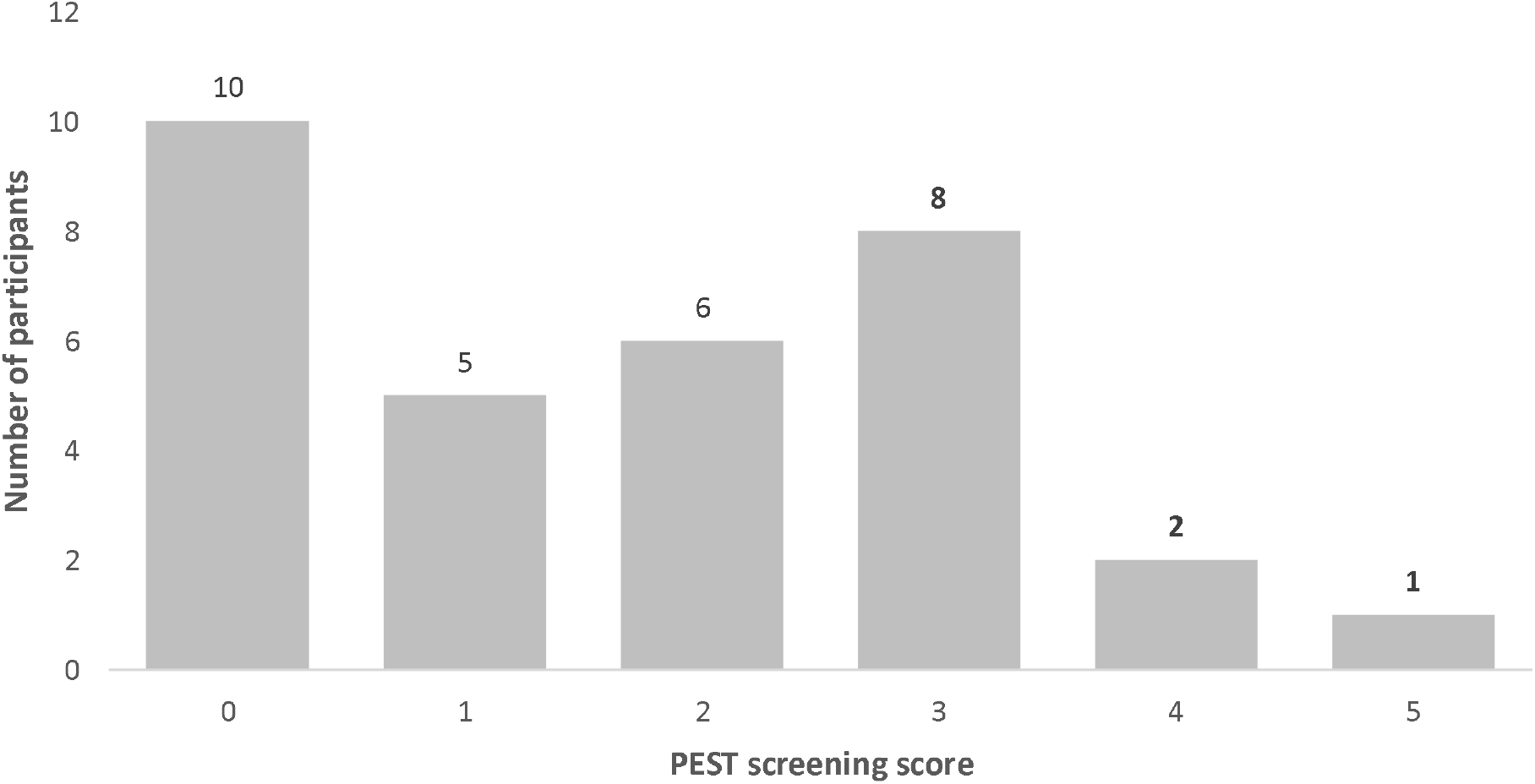
PEST screening scores by patient count

As shown in Figure 3, a total of 10 patients scored 0, 5 patients scored 1, 6 patients scored 2, 8 patients scored 3, 2 patients scored 4, and 1 patient scored 5. PEST screening services administered by pharmacists in a specialty setting identified 11 of 32 patients (34%) who scored a 3 or higher and may benefit from a referral to a rheumatologist for further assessment of psoriatic arthritis activity. Out of the 11 patients who screened positive, 8 patients (73%) continued on their current biologic treatment (ustekinumab n=4, adalimumab n=2, risankizumab n=2). All three of these agents are FDA approved for treatment in both plaque psoriasis and psoriatic arthritis and remain appropriate options for patients who screened positive for psoriatic arthritis activity. Two patients had therapeutic switches, secukinumab to ixekizumab and guselkumab to Remicade (infliximab). One patient discontinued drug treatment with ixekizymab due to loss of insurance. Of note, one patient had a diagnosis of psoriatic arthritis added to their medical history and mentioned within medical charts after completing the PEST screen. This observation suggests a discussion of psoriatic arthritis activity with their medical provider as a result of the assessment administered by the specialty pharmacy.

## DISCUSSION

To our knowledge, there are no known studies documenting PEST screening initiatives by pharmacists in the specialty setting and only one article which describes patient outcomes using the RAPID3 in those with rheumatoid arthritis. This single, retrospective study documented the applicability of a RAPID3 assessment built-into a specialty pharmacy’s dispensing system to monitor a total of 265 patients with moderate to high rheumatoid arthritis activity over a nine-month period. Researchers were able to conclude that technology-enabled clinical support has the potential to improve compliance with practice guidelines. This study had a technology-based focus to demonstrate how specialty pharmacists can expand their role in monitoring and enhancing outcomes by utilizing validated technological instruments.^23^

Although the impact of clinical pharmacist screening services is well documented in many other chronic conditions such as peripheral artery disease, COPD, and osteoarthritis, the capability and importance in the specialty setting has yet to be established. The study aimed to describe the impact of pharmacist-administered screenings in rheumatoid arthritis patients to depict outcome monitoring through disease severity and therapeutic switches. RAPID3 data showed that disease severity category significantly improved from baseline to final assessment, with 21 patients responding to treatment and 11 patients having therapeutic switches. These results suggest the clinical advantage of specialty pharmacists working in collaboration with providers to increase the number of responders and maximize drug therapy through screening initiatives. Non-responders comprised 12.2% of the RAPID3 study population, highlighting patients who may benefit from referral to their provider for reevaluation of their therapeutic plan. As shown in this study, these assessments enhance patient care by identifying persons who are not responding to therapy as well as identifying persons who are at risk for more advanced disease. An additional takeaway from this data is the recognition that screening and monitoring of complex inflammatory diseases can be fully incorporated to fit within routine workflow of a clinical specialty pharmacist.

In patients with psoriasis, the primary goal was to describe the utilization of a screening for psoriatic arthritis and the resulting opportunity for referral to medical providers. Pharmacists in the specialty setting were able to identify 11 patients (34%) with psoriatic arthritis activity. This demonstrates the ability of the assessments to highlight opportunities for therapy changes and serve as a tool for outcome monitoring. Furthermore, the screenings call attention to biologic indications to ensure the optimization of therapies that treat comorbid inflammatory conditions, such as psoriasis and psoriatic arthritis, based on FDA approvals. The PEST screening results further demonstrates the value of clinical pharmacists in the specialty setting and their essential role in improving patient outcomes while working within a health care team.

## LIMITATIONS

The primary limitations to this study are sample size, geographic location, and length of study duration. The study was limited to 32 participants in the PEST screening group over a three-month period and 127 patients in the RAPID3 screening group over a thirteen-month period. The study population represented patients filling at a single specialty pharmacy in one regional location.

## CONCLUSION

The study demonstrates the impact of specialty pharmacist screening services in describing disease activity in rheumatoid arthritis patients using the RAPID3 and suggesting need for further referral in psoriasis patients without confirmed psoriatic arthritis using the PEST tool. Clinical pharmacists in the specialty setting are an essential part of improving patient outcomes, maximizing therapeutic regimens, and working in collaboration with other healthcare professionals.

## Data Availability

All data produced in the present work are contained in the manuscript.

